# Evaluation of a Next Generation Sequencing Assay for Hepatitis B Antiviral Drug Resistance on the Oxford Nanopore System

**DOI:** 10.1101/2024.10.01.24314730

**Authors:** Michael Payne, Gordon Ritchie, Tanya Lawson, Matthew Young, Willson Jang, Aleksandra Stefanovic, Marc G. Romney, Nancy Matic, Christopher F. Lowe

**Affiliations:** Division of Medical Microbiology and Virology, St. Paul’s Hospital, Providence Health Care, Vancouver, British Columbia, Canada; Department of Pathology and Laboratory Medicine, University of British Columbia, Vancouver, British Columbia, Canada

**Keywords:** Hepatitis B virus, Viral resistance testing, Next generation sequencing, Molecular methods

## Abstract

**Background:** Next-generation sequencing (NGS) for Hepatitis B virus (HBV) antiviral resistance (AVR) testing is a highly sensitive diagnostic method, able to detect low-level mutant subpopulations. Our clinical virology laboratory previously transitioned from DNA hybridization (INNO-LiPA) to NGS, initially with the GS Junior System and subsequently the MiSeq. The Oxford Nanopore Technology (ONT) sequencing system was evaluated for HBV resistance testing, with regards to sequencing accuracy and turn-around time.

**Methods:** We performed amplicon sequencing of the HBV polymerase gene from patient plasma and external quality assessment (EQA) samples on the MiSeq Reagent Nano Kit v2 and GridION ONT with R10.4.1 flowcells. Mutational analysis and genotyping were performed by DeepChekAssay-HBV (version 2.0).

**Results:** A total of 49 patient samples and 15 EQA samples were tested on both the MiSeq and ONT. There was high agreement for both patient and EQA samples between the MiSeq and ONT systems, with regards to total drug resistance mutations detected and total patient sample agreement, 68/70 (97%) and 47/49 (96%), respectively.

**Conclusion:** The ONT NGS platform provided accurate HBV AVR results, with improved turn- around times. Sequencing error rates at AVR codons were below 1%.

## 1. Background

Hepatitis B virus (HBV) is an important cause of chronic hepatitis and hepatocellular carcinoma (HCC) worldwide. Globally, there are an estimated 296 million people living with HBV, along 1.1. million deaths annually (1). Treatment of chronic HBV can decrease the risk of progression to cirrhosis and HCC. Identification of the HBV genotype, along with antiviral resistance (AVR) mutations, can help inform the management of chronic HBV infection (2–4). Newer antiviral agents (Entecavir and Tenofovir) have a higher barrier to AVR mutations and viral breakthrough; however, AVR testing is useful to monitor for emergence of mutations associated with decreased virologic response, particularly for patients with previous exposure to HBV antiviral therapies (3, 5).

HBV AVR line probe (LiPA) testing is limited by its inability to detect novel mutations, hybridization failures, and decreased sensitivity with low viral loads (<1000 IU/mL) (6, 7). In 2014, testing in our clinical laboratory was transitioned to next-generation sequencing (NGS) with the GS Junior System (454 Life Sciences, Branford, CT) and in 2017, the MiSeq (Illumina) platform. MiSeq offered increased sequencing depth; however, a shorter read length did not include all AVR mutation sites (8).

The Oxford Nanopore Technology (ONT) sequencing system (GridION ONT) with longer read capability and updated R10.4.1 flowcells has reported improved error rates, allowing for more accurate detection of variant subpopulations (9). In addition, this technology has less hands-on library preparation and sequencing run time, allowing for improved workflow in clinical laboratories. Our study aim was to optimize HBV AVR testing using NGS on the ONT platform, along with validation of this test for clinical use.

## 2. Materials and Methods

We used guidance from molecular testing references and draft recommendations from the US FDA to design this NGS validation study (10–12). Parameters evaluated included: Limit of detection, inclusivity/accuracy, reproducibility/precision and cross-contamination. DNA was extracted from samples using the MagNA Pure LC 2.0 (Roche Diagnostics, Mannheim, Germany) for MiSeq and the MagNA Pure 24 (Roche) for ONT. For both ONT and MiSeq, PCR primers amplified an 820 base pair (bp) region of the HBV polymerase gene on the Roche LightCycler® 480. Primer sequence (5’ → 3’) forward (ILF) CGT GGT GGA CTT CTC TCA ATT TTC and reverse (ILR) AGA AAG GCC TTG TAA GTT GGC GA. For MiSeq, the second nested PCR reaction mixture consisted of the first round PCR product, and fusion indexed and barcoded primers. The nested PCR final amplicon was 364 bp. ONT AVR sequencing was performed using the GridION with R10.4.1 flow cells and the SQK-NBD114.24 library kit (Oxford Nanopore Technologies). Guppy (v. 6.4.6) was utilized for sequence base-calling. For MiSeq and ONT, mutational analysis and genotyping of FASTQ files was performed with DeepChek® HBV v.2.0 (ABL SA Group). Interpretation of clinically significant AVR mutations was based on the 2017 EASL Clinical Practice Guidelines (2).

The 2nd World Health Organization (WHO) International Standard for Hepatitis B Virus DNA was used for evaluation of the limit of detection for the initial HBV PCR. Serial dilutions of the WHO Standard were tested with 5 replicates at: 1000, 500, 250, 125, 62.5, and 30 IU/mL. Probit regression was performed to determine limit of detection.

A total of 49 patient samples and 15 external quality assurance (EQA) samples (QCMD, Glasgow, United Kingdom) were selected for AVR testing by NGS, to include a broad range of AVR mutations, at varying viral loads and percentage mutation frequency. There were 51 patient and 35 EQA samples available for genotype testing. The ATCC 45020D plasmid DNA was utilized as a control in all runs to determine error rates for base calling.

To evaluate reproducibility/precision, a well characterized patient sample (genotype C; viral load of 9.9 × 10^8^ IU/mL) was utilized, with mutations at approximately 70%, 51%, 26%, and 7%, at codons M204I, V173L, L180M, and T184S, respectively. The patient sample was diluted in Basematrix plasma (Seracare, Milford, MA) to 10,000, 4000, 1000, 250 and 125 IU/mL. These were tested in 3-5 total replicates over 2 separate NGS runs to determine the reproducibility and sensitivity of NGS at detecting different percent mutations at various viral loads. In order to assess cross-contamination risk, each run contained a negative control.

## 3. Results

There was a high agreement rate between MiSeq and ONT with regards to total mutations detected and patient sample agreement, 68/70 (97%) and 47/49 (96%), respectively. All EASL mutations were included in the study, with the exceptions of M250V, N236T and T184G (Table 1) (2).

**Table 1.**
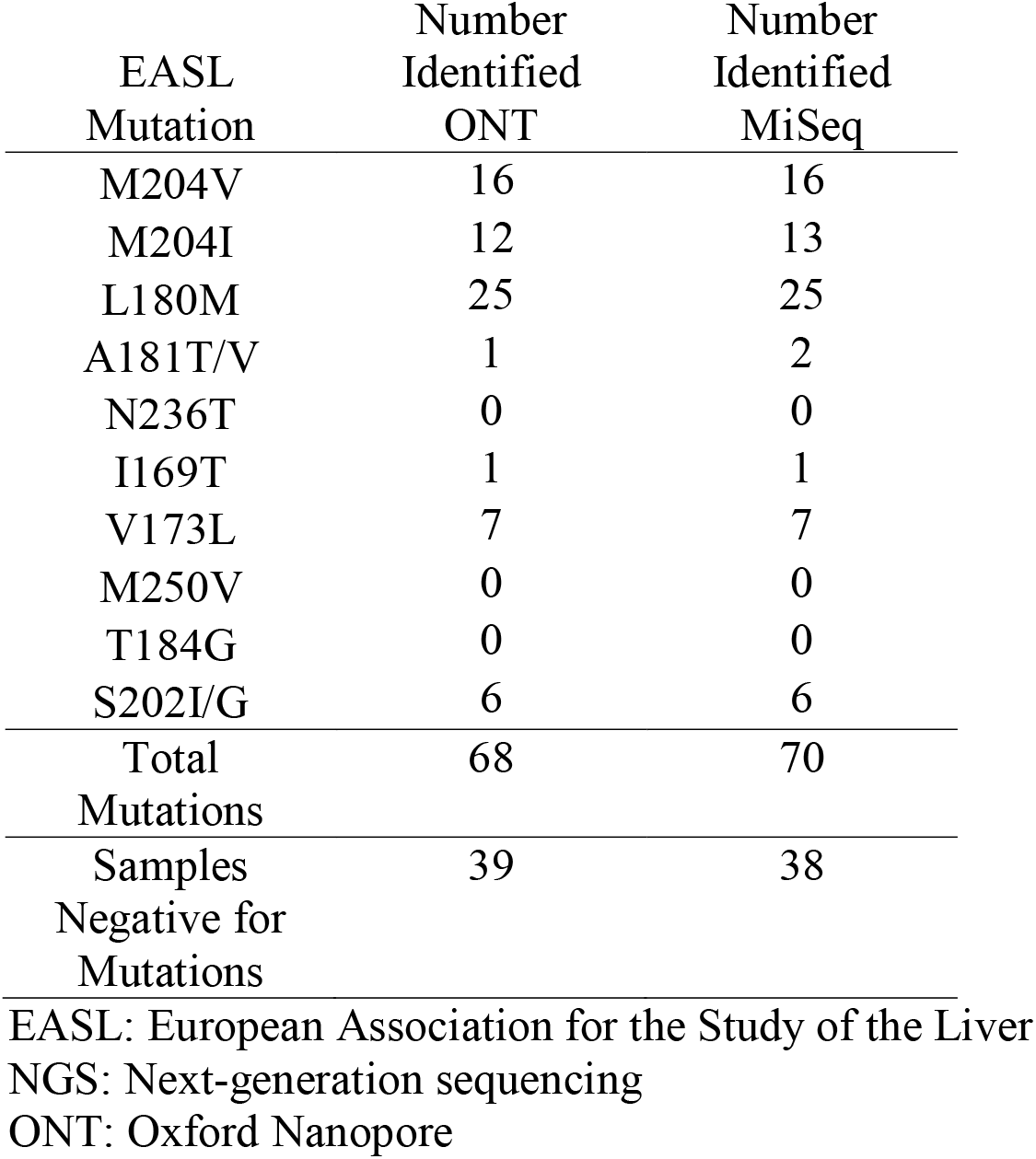
Hepatitis B Antiviral Resistance Mutations Identified by Next Generation Sequencing.

There were two patient samples with a mutation detected only by MiSeq (Table 2). The discrepant mutation results were found with low viral load samples (<2000 IU/mL). For one sample, the mutant subpopulation was detected at a low frequency (4.3%, M204I) only on MiSeq and would not have changed the genotypic resistance interpretation as a M204V mutation was also detected at a high frequency on both MiSeq and ONT platforms. For the second sample, the A181T mutation was found on MiSeq at a higher frequency (38%). QCMD samples with known HBV AVR mutations were tested by MiSeq and ONT. There was 80% (12/15) agreement for both platforms, with 3 samples having a L80I/V mutation identified only by the ONT platform. This is a known limitation of the MiSeq assay, as the sequenced amplicon is shorter and does not cover that particular mutation.

**Table 2.**
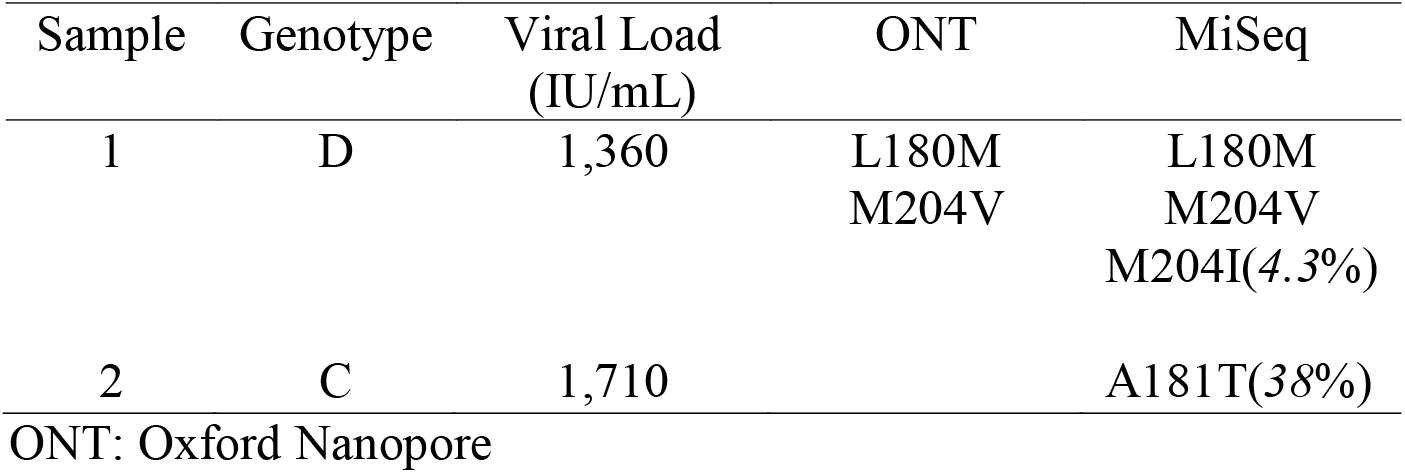
Discordant Result Analysis.

| Sample | Genotype | Viral Load<br>(IU/mL) | ONT | MiSeq |
| --- | --- | --- | --- | --- |
| 1 | D | 1,360 | L180M<br>M204V | L180M<br>M204V<br>M204I(4.3%) |
| 2 | C | 1,710 |  | A181T(38%) |

ONT: Oxford Nanopore

There were 35 EQA samples tested with the following genotypes: A, B, C, D, F. All were identified accurately by MiSeq and ONT. A total of 51 patient samples were tested for genotype by both MiSeq and ONT, and all results were concordant. These were composed of HBV genotypes: A (9), B (12), C (22), D (5), E (2), and A/G (1).

ONT showed reproducible sequence variant detection for replicates with high viral loads and/or high-level sequence variants (Table 3). Sequence variant detection showed increased variability with low viral loads samples (<1000 IU/mL), particularly with low-level frequency subpopulations (<10%). The limit of detection for the initial PCR was 119 IU/mL. From the plasmid control, average base calling error rates at AVR codons were: V173 (0.74%), L180 (0.84%), M204 (0.36%) and M250 (0.42%). There was no evidence of sample cross-contamination risk, with each runs negative control testing negative.

**Table 3.** Reproducibility in minority variant analysis with decreasing viral loads.

| Variant | Viral Load<br>(IU/mL) | ONT |  |
| --- | --- | --- | --- |
|  |  | Mean (%) | Coefficient of variation |
| M204I | 10000 | 70.3 | 7.8 |
|  | 4000 | 67.7 | 10.8 |
|  | 1000 | 72.0 | 17.6 |
|  | 250 | 89.3 | 17.1 |
|  | 125 | 41.6 | 115.8 |
| V173L | 10000 | 50.5 | 7.5 |
|  | 4000 | 48.2 | 10.1 |
|  | 1000 | 61.4 | 22.3 |
|  | 250 | 45.2 | 84.5 |
|  | 125 | 24.0 | 195.3 |
| L180M | 10000 | 25.7 | 21.6 |
|  | 4000 | 31.7 | 24.8 |
|  | 1000 | 25.0 | 48.3 |
|  | 250 | 7.9 | 190.7 |
|  | 125 | 54.5 | 86.5 |
| T184S | 10000 | 6.5 | 16.4 |
|  | 4000 | 10.3 | 92.7 |
|  | 1000 | 1.1 | 120.9 |
|  | 250 | 2.5 | 95.2 |
|  | 125 | 0.3 | 15.4 |

ONT: Oxford Nanopore

The ONT assay PCR and library preparation time was approximately 30% less than the MiSeq assay (5 versus 7 hours for one sample with controls); however, the time required for library preparation is also dependent on the number of samples included with each sequencing run. Sequencing time is markedly reduced with the ONT platform: approximately 15-30 minutes for ONT as compared to 23 hours for MiSeq assay. The time required for bioinformatic analysis for each sample was comparable between the assays (∼15 minutes).

## 4. Discussion

This study describes the clinical validation of a NGS HBV AVR assay using the ONT platform. Results for ONT were highly concordant with MiSeq, with 100% (51/51) accuracy for genotype and 96% (47/49) accuracy for AVR testing on patient samples. Two samples had an additional AVR mutation detected by MiSeq which were not detected by ONT. These additional mutations were detected only on MiSeq from samples with low viral loads (<2000 IU/mL). The increased detection of low-level subpopulations by the MiSeq assay could be a result of the increased read depth per run with the MiSeq platform (∼12,500) as compared to ONT (∼8000). Additionally, the MiSeq assay used a more sensitive nested PCR for amplicon sequencing, which may explain improved detection of AVR mutations in low viral load samples. The nested PCR step was discontinued for the ONT assay, as it was more labour intensive, and had increased risks for sample cross-contamination. In addition, for samples with HBV viral load <500 IU/mL, our laboratory routinely reports a qualifying comment regarding decreased sensitivity for detecting mutant subpopulations of <10%, which is consistent with the reproducibility results from Table 3. ONT was able to improve PCR/library preparation and sequencing TAT, which has been previously demonstrated (13). Using the 45020D plasmid DNA control, which has a known sequence, ONT sequencing error rates were found to be less than 1% (0.36-0.84%). This is consistent with error rates reported for the R10.4.1 flow cells (9). Sequencing error rates are minimized by further quality filtering by Q score and forward/reverse read bias performed during bioinformatic analysis. This allows for confidence in reporting mutations found at a frequency of 3.5% or greater.

Although resistance to antiviral agents is uncommon with entecavir and tenofovir, particularly in treatment-naïve patients, accessibility to HBV AVR is required for clinical follow up of patients with virological breakthrough to guide treatment (2–4). While *de novo* resistance to entecavir is rare, resistance can develop over time with chronic entecavir treatment: 0.9% at ≥5 years for treatment naïve and 20.1% for nucleos/tide analogue experienced (14). Limited data exist regarding tenofovir resistance, though suspected mutations have been reported which were not described in the EASL guidance (2, 5). In addition to tenofovir, novel antiviral agents aiming for functional cure are in the pipeline, and AVR sequencing will be needed to provide ongoing surveillance for any potential novel mutations associated with drug resistance (15, 16).

There are practical barriers to the introduction of NGS in clinical laboratories, including cost, technical expertise and data analysis/interpretation (17, 18). As a clinical laboratory without a dedicated bioinformatician, we utilized a commercial platform to analyze FASTQ sequencing data and to remove the barrier relating to bioinformatics. Laboratories can also consider other commercial bioinformatic tools, which may increasingly become available as more laboratories launch NGS testing programs (19). With developing infrastructure and demand for clinical sequencing, laboratories may also consider the feasibility of in-house developed bioinformatic pipelines to enable customization of sequence interpretation, particularly for novel drugs/mutations (20).

Limitations of our study include, despite attempts to include a broad array of AVR mutations and genotypes (patient and EQA samples), less frequently encountered AVR mutations and genotypes could not be included. In addition, the clinical relevance of identifying low-level subpopulations of AVR mutations through NGS is still unclear, and further research is required to determine relevant thresholds for identification of low-level subpopulations.

In conclusion, a single real-time PCR followed by NGS with the ONT platform was found to be highly accurate for the detection of HBV AVR mutations and HBV genotype testing in a clinical laboratory. This was consistent across a wide variety of HBV genotypes and resistance mutations. Benefits of the ONT HBV assay included increased read length, decreased sample processing time, and improved TAT.

## Data Availability

All data produced in the present study are available upon reasonable request to the authors.

## Authorship

Conceptualization, MP, GR, NM, CFL; Data curation, MP, GR, TL, MY, WJ; Formal analysis, MP, GR; Sequencing and analysis, GR; Funding acquisition, N/A; Investigation, MP, GR, AS, MGR, NM, CFL; Methodology, GR, TL, MY; Project administration, WJ, MGR, NM, CFL; Supervision, NM, CFL; Visualization, MP; Writing – original draft, MP; Writing – review & editing, MP, GR, TL, MY, WJ, AS, MGR, NM, CFL.

